# Aspirin reduces the mortality risk of Acute Respiratory Failure: an observational study using the MIMIC IV database

**DOI:** 10.1101/2024.11.27.24318046

**Authors:** Zhenhong Jiang, Shijin Lv, Guohu Zhang, Zengyan Fu

**Author notes:** Le Jiang(corresponding author) from Affiliated Hospital of Hangzhou Normal University.

## Abstract

**Background:** Acute Respiratory Failure (ARF) is a serious complication of various diseases, characterized by a high mortality rate. Aspirin influences cyclooxygenase, which have a crucial role in inflammation, blood clotting, and immune system modulation. ARF is characterized by an uncontrolled inflammatory and pro-coagulant response, but aspirin can mitigate this inflammatory response by inhibiting platelet function, potentially leading to improved outcomes.Numerous studies have produced conflicting data concerning the impact of aspirin on individuals suffering from Acute Respiratory Failure. We performed an analysis of the MIMIC IV database to explore the association between aspirin use and the outcomes in ARF patients, as well as to ascertain the optimal dosing regimen for aspirin treatment.

**Materials and methods:** ARF patients’ clinical data were extracted from MIMICIV2.2. Propensity score matching was utilized to ensure comparability of baseline characteristics between the group receiving aspirin and the group not receiving aspirin. Subsequently, the link between aspirin and patient death was examined through the application of Kaplan-Meier estimations and Cox proportional hazard regression analyses.

**Results:** We identified a cohort of 6,663 individuals suffering from ARF from the MIMIC IV database.Following propensity score matching in a sample of 4,008 participants, multivariate Cox proportional hazards analysis revealed a lower hazard of dying within 90 days for those in the aspirin group versus the non-users group (adjusted Hazard Ratio: 0.723; 95% Confidence Interval: 0.652 to 0.802). Additionally, the Kaplan-Meier survival curves indicated that the 90-day survival rate was higher among aspirin users compared to non-users (log-rank test p< 0.001). And the median duration of survival for patients undergoing aspirin therapy was considerably extended compared to those who did not receive it, amounting to 15.60 days versus 10.36 days, respectively In the aspirin group, the median ICU stay length was longer than non-users group (6.93 days vs. 6.08 days, p <0.001).Patients in the aspirin use group had a significantly shorter duration of mechanical ventilation compared to those in the non-aspirin use group(54.95 days vs. 58.00 days, p <0.001).

**Conclusion:** Aspirin may reduce the 30-day or 90-day mortality risk in ARF patients and also shorten the duration of mechanical ventilation. Aspirin could be an effective medication in the treatment of ARF patients.

## 1 introduction

Acute respiratory failure (ARF) is a critical condition characterized by the abrupt inability of the lungs to maintain adequate oxygenation and ventilation, leading to life-threatening respiratory distress^1^. The etiology of ARF is diverse, including pneumonia, sepsis, trauma, and other non-infectious causes such as pancreatitis or drug-induced lung injury^2^. As a life-threatening condition, an estimation suggests that in the United States, close to 2 million individuals suffering from acute respiratory failure are admitted to hospitals every year, accruing expenses that surpass $50 billion annually^3^. Around 50% of individuals with acute respiratory failure need invasive mechanical ventilation, and the proportion of these patients who do not survive their hospital stay is over 20%^3^.Current treatment strategies of ARF primarily focuses on supportive care, which may involve mechanical ventilation, oxygen therapy, and hemodynamic support^4,5^. Despite advances in critical care, there remains a lack of targeted pharmacological therapies that directly address the underlying pathophysiology of ARF. This condition often involves complex interactions between the coagulation and inflammatory systems, with the endothelium playing a central role in the disease’s progression^6^. In the pathophysiological progression of ARF, the normal antithrombotic and anti-inflammatory phenotype of endothelial cells can shift towards a prothrombotic and proinflammatory state. This transition is marked by an increase in procoagulant activity within the pulmonary vascular bed and a decrease in fibrinolytic capacity, leading to fibrin deposition and alveolar damage^7,8^. While fibrin deposition may initially support gas exchange and tissue repair, it can also contribute to further pulmonary dysfunction and fibrosis if unchecked^5^. As research continues, there is a growing interest in identifying novel therapeutic targets and biomarkers that could improve outcomes in patients with ARF.

Aspirin, a ubiquitously utilized medication, is renowned for its fever-reducing, pain-relieving, and inflammation-lowering attributes , owing to its action on cyclooxygenases—enzymes pivotal in inflammation, blood clotting, and immune system modulation^9,10^. Emerging studies have expanded our understanding of aspirin’s versatility, demonstrating its efficacy in inhibiting platelet aggregation to mitigate the likelihood of subsequent heart attacks in cardiovascular disease sufferers, as well as its potential role in colorectal cancer prevention^11^. According to relevant reports, aspirin has the capability to suppress platelet activity and dampen inflammation, leading to enhanced outcomes^10^. Several studies have reported aspirin as a promising treatment option for ARF^9,12–15^. However, Daryl J et al. have reported that the use of aspirin compared with placebo did not reduce the risk of ARDS at 7 days^15^. Philip et al. have discovered that aspirin did not improve OI or other physiological outcomes^12^. These research outcomes have presented conflicting findings on the impact of aspirin therapy in individuals suffering from Acute Respiratory Failure (ARF). And aspirin is a special antiplatelet drug because different dosage of aspirin has different function ^16^. Low-dose aspirin, typically ranging from 75 to 100 milligrams per day, is recognized for its ability to inhibit platelet aggregation, while higher doses, exceeding 300 milligrams per day, not only affect platelet function but also possess anti-inflammatory properties. However, prior research has not specifically considered the most effective dosage of aspirin for patients with ARF^10,17^. Therefore, we carried out an examination of the MIMIC IV database to explore the association between the use of aspirin and the clinical outcomes in patients suffering from ARF. Additionally, our aim was to ascertain the optimal dosing regimen for aspirin treatment within this patient population.

## 2 Materials and methods

MIMIC-IV, the fourth iteration of the Medical Information Mart for Intensive Care, is a comprehensive, single-site database that offers open access to a vast trove of information. Specifically, it encompasses records from over 730,000 ICU admissions at the Beth Israel Deaconess Medical Center in the U.S., with data spanning from 2008 through 2019^18^. This resource can be found at https://physionet.org/content/mimiciv/2.2/.Author Zhenghong Jiang retrieved clinical data, encompassing patient demographics, laboratory findings, and medication records, from the MIMIC database (with certification number: 12918901). Utilization of this database received endorsements from the institutional review boards at MIT and BIDMC. This project was conducted in accordance with the ethical guidelines outlined in the Declaration of Helsinki. Given the de-identified status of the participants and the structured data in the database, further ethical committee clearance was not deemed necessary for this investigation.

## 3 Inclusion and exclusion criteria

Select patients who met the diagnosis of acute respiratory failure based on ICD codes and included them in this study. The exclusion criteria were: 1) ICU stay duration < 48 h (discharged or death); 2) Patients aged <18 years. The ARF patients who received aspirin in the ICU were compared to those who did not.

### 3.1 Data collection

Using the Navigate Premium tool, version 16, Structured Query Language (SQL) queries were employed to retrieve information pertaining to patients from the MIMIC-IV database(version 2.2). The following ARF comorbidities were recorded: cancer, diabetes mellitus, sepsis, COPD, hypertension, acute pancreatitis and renal disease. Subsequently, the laboratory indexes within the first day of ICU admission were extracted, including hemoglobin, white blood cells, platelets, prothrombin time, and Oxygenation index. SAPS II scores, SOFA scores and OASIS scores were calculated at ICU admission. The following treatment information was noted: use of mechanical breathing, vasopressors, renal replacement therapy (RRT) and continuous renal replacement therapy(CRRT).

### 3.2 Primary outcome and secondary outcomes

The primary outcome of this study was the 90-day mortality. The secondary outcomes included 30-day mortality and ICU stay duration.

### 3.3 Statistical analysis

The variables in our study had missing values, so we would remove them. Patients diagnosed with ARF who were administered aspirin during their stay in lCU were categorized into the experimental cohort. Conversely, ARF patients who did not receive aspirin therapy comprised the control cohort. For continuous variables that did not follow a normal distribution, the Mann-Whitney U test was applied, and the results were presented using medians accompanied by interquartile ranges. Categorical variables between the two groups were depicted by counts and their respective proportions, and differences were evaluated utilizing the chi-squared test. All statistical analyses were carried out employing the SPSS Statistics software, version 26.0. A p-value of 0.05 or below was considered indicative of statistical significance.

To minimize the disparity in baseline characteristics between the two cohorts, a propensity score matching technique was employed with a caliper width set at 0.02. This method helps to create more comparable groups by matching participants based on their probability of receiving the treatment or exposure, thereby reducing selection bias and confounding factors.

Through the application of Kaplan-Meier estimators, graphical representations demonstrated the proportions of ARF patients surviving up to 30 days and 90 days, further highlighting the midpoint of survival times for the cohort. To evaluate the effect of aspirin treatment on the risk of dying within 30 days and 90 days in patients with acute respiratory failure, we utilized the Cox proportional-hazards model to calculate the hazard ratios and corresponding confidence intervals. Further, acute respiratory failure patients were grouped by demographic and clinical criteria, encompassing age, gender, race, comorbid conditions, severity assessment scores (SAPS II, SOFA, OASIS), and interventions such as renal replacement therapy (RRT), continuous renal replacement therapy (CRRT), mechanical breathing support, and use of vasoactive agents. Individual subgroup analyses were performed to ascertain hazard ratios and the corresponding 95% confidence intervals for each category.

## 4 Results

### 4.1 Patient characteristics

The process by which patients were chosen for inclusion in this current research is depicted in Figure 1. In summary, a total of 6,663 patients diagnosed with ARF satisfied the predetermined inclusion criteria for the study. The comprehensive clinical data pertaining to these 6,663 ARF patients, which includes the initiation and cessation timelines of aspirin therapy, as well as the dosage administered, is provided in Supplementary Table S2. It is pertinent to note that the information regarding aspirin treatment is absent for the group of patients who did not receive aspirin (the non-user aspirin group). In our study, the most common dose of aspirin is 81 mg/d (2,286 patients).There are 462 patients using high-dose aspirin (>300 mg/d).Our investigation is a retrospective analysis, which inherently carries the potential for confounding variables to influence the outcomes significantly. Propensity score matching (PSM) is a statistical technique widely employed to mitigate the impact of confounding variables. Essentially, it pairs individuals from the treatment group with those from the control group who share comparable baseline characteristics. 6,663 patients with ARF were extracted from the MIMIC IV database. Through the application of Propensity Score Matching (PSM), we successfully paired 4,008 individuals, effectively reducing the influence of confounding variables. Post-matching, the primary determinant under scrutiny became the administration of aspirin, with the effects of other potential confounders substantially diminished. This allowed for a more focused analysis on how aspirin use might correlate with the outcomes being studied, providing a clearer picture of its potential impacts.It displays the clinical information of ARF patients of ARF patients in the two groups before or after Propensity score matching (PSM), in the Table 1. During their stay in the ICU, 2,813 of the ARF patients were administered aspirin, while 3,850 of the ARF patients did not receive aspirin therapy. Among the ARF patients, 920 cases (13.8%) were diagnosed with cancer, 2,159 cases (32.4%) had diabetes, and 2,527 cases (37.9%) suffered from congestive heart failure. A total of 1,276 patients (19.2%) had experienced myocardial infarction, and the percentage of patients with sepsis stood at 87.9% (5,860 cases). Additionally, 2,251 cases (33.8%) had Chronic Obstructive Pulmonary Disease (COPD). Moreover, the proportions of patients with hypertension, acute pancreatitis, and renal disease were 53.5% (3,565 cases), 3.5% (233 cases), and 25.3% (1,687 cases), respectively.Compared to subjects who did not receive aspirin, patients who were on aspirin therapy had higher prevalence rates of diabetes (16.20%), congestive heart failure (54.90%), myocardial infarction (34.10%), COPD (38.30%), hypertension (60.40%), and renal disease (33.70%). Furthermore, the average age in the group receiving aspirin was greater than in the non-user group (Table 1).

**FIGURE 1.**
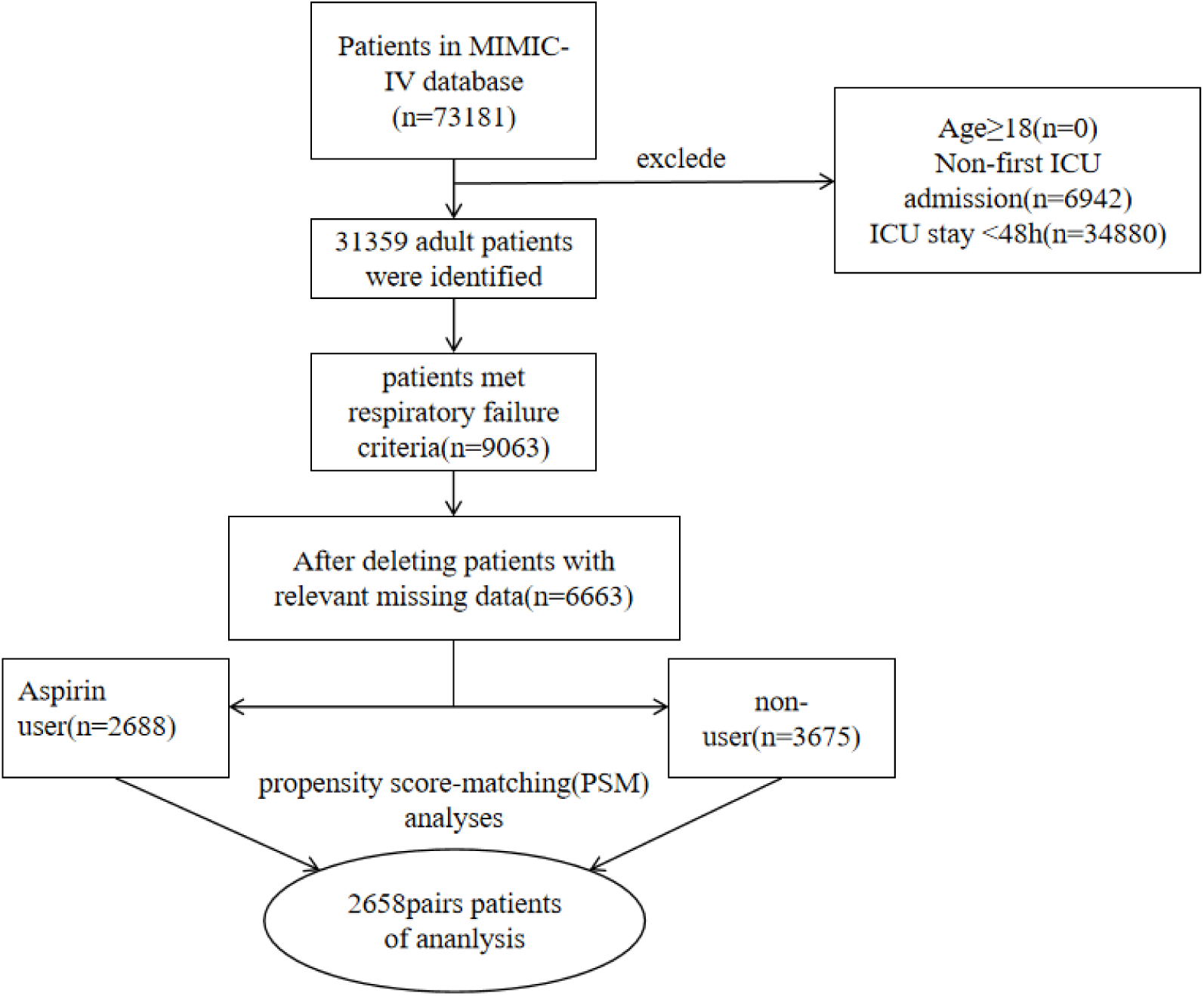
Flow diagram of the research.

**TABLE 1.**
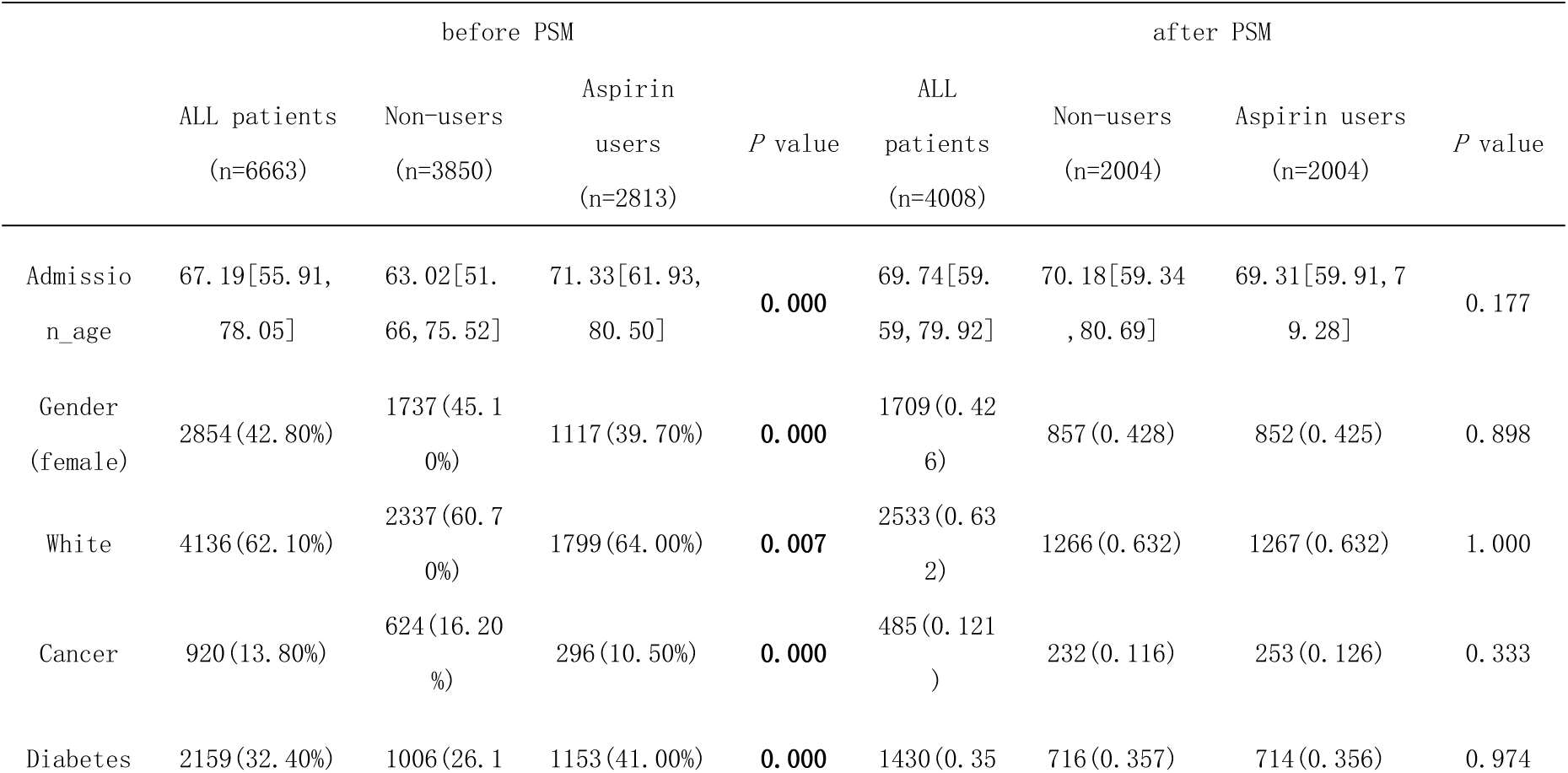
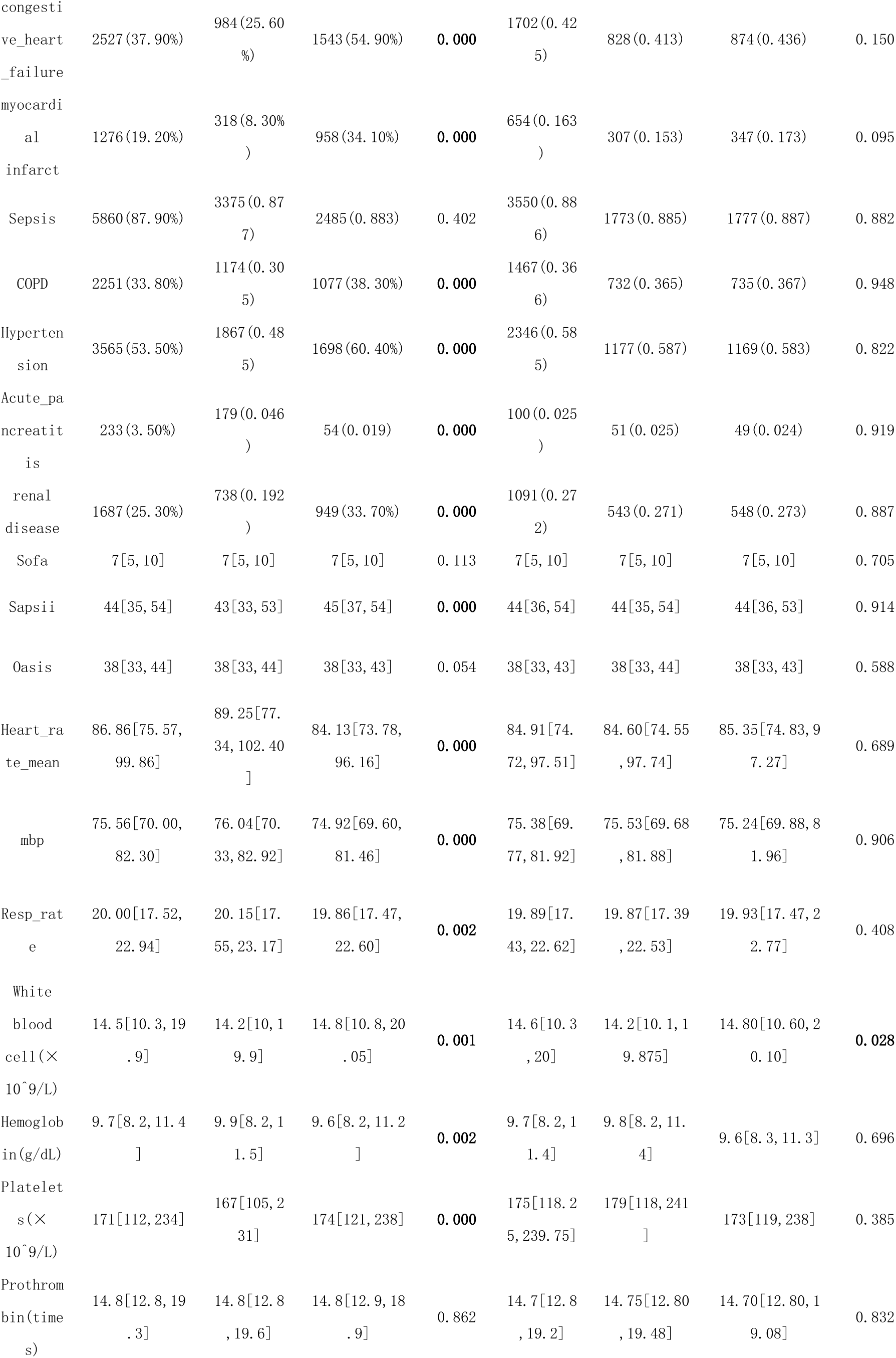
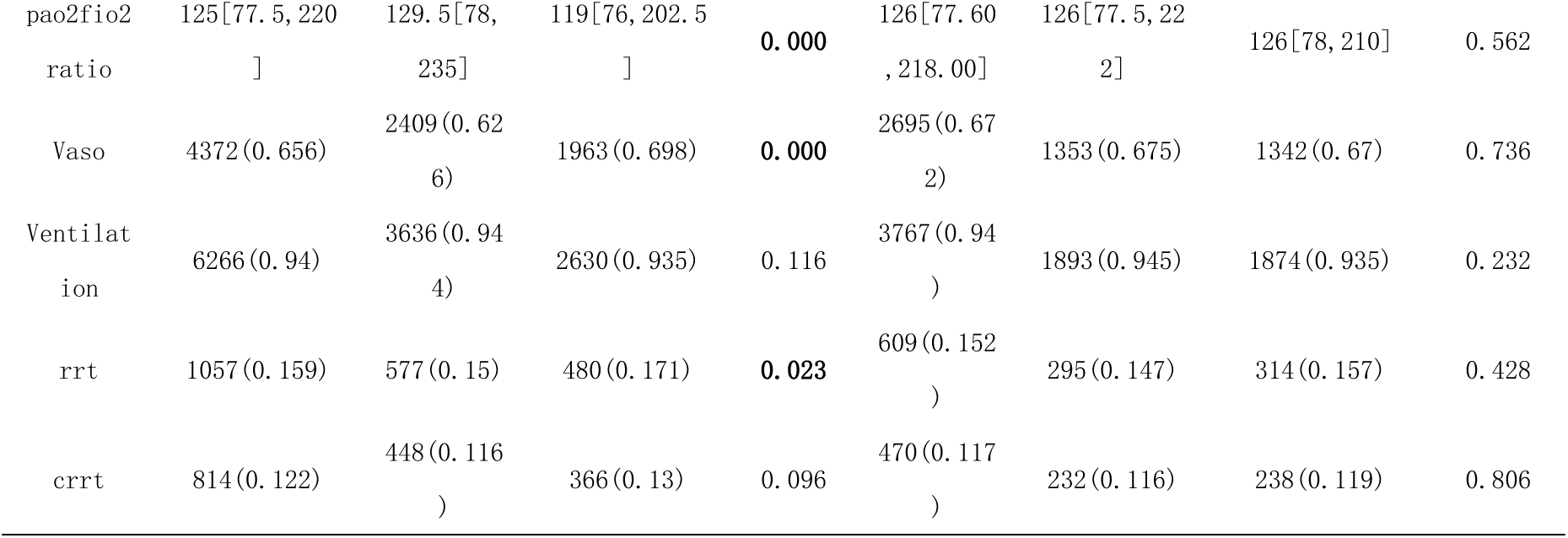
Clinical information of ARF patients before PSM and after PSM.

### 4.2 Association between aspirin and mortality outcomes

ARF patients were categorized into two groups based on whether they received aspirin during their hospitalization. Compared to the non-user group, the aspirin group demonstrated lower risks of death at both 30 days and 90 days [Hazard Ratio (HR): 0.815; 95% Confidence Interval (CI): 0.724-0.918; HR: 0.723; 95% CI: 0.652-0.802], as revealed by univariate Cox regression analysis (Table 2). The Kaplan-Meier curves for 30-day survival are depicted in Figure 2, and for 90-day survival in Figure 3. According to the Kaplan-Meier survival analysis, the 30-day and 90-day survival rates were significantly higher in the aspirin user group than in the non-user group (Log-rank test: p-value < 0.001). And patients who received aspirin treatment had a notably longer median survival time at 90 days compared to those who did not receive the treatment (15.60 days vs. 10.36 days) (Table 2).We analyzed the relationship between aspirin and mortality in ARF patients, along with other potential predictors of survival, using Cox regression. The aspirin group showed a lower risk of death at 30 days and 90 days compared to the non-user group (Adjusted HR=0.757, 95% CI: 0.669-0.858; Adjusted HR=0.699; 95% CI: 0.628-0.778), as determined by multivariate Cox proportional hazards analysis (Table 2).

**TABLE 2.** Survival outcomes of the aspirin group and the non-users group

|  | Group | No. events/No. All patients | Median survival time (95%CI) | Log rank test value | Hazard ratio (95%CI) | Adjusted hazard Ratio (95%CI) |
| --- | --- | --- | --- | --- | --- | --- |
| 30-day mortality | Non-users | 688/2044 | 8.22(4.24,14.47) | <0.001 | HR=0.815 (0.724,0.918) | Adjusted HR=0.757(0.669,0.858) |
|  | Aspirin-users | 451/2044 | 10.48(6.28,17.07) |  |  |  |
| 90-day mortality | Non-users | 830/2004 | 10.36 (5.08,21.29) | <0.001 | HR=0.723(0.625,0.802) | Adjusted HR=0.699(0.628,0.778) |
|  | Aspirin-users | 637/2004 | 15.60 (8.02,34.20) |  |  |  |

**FIGURE 2.**
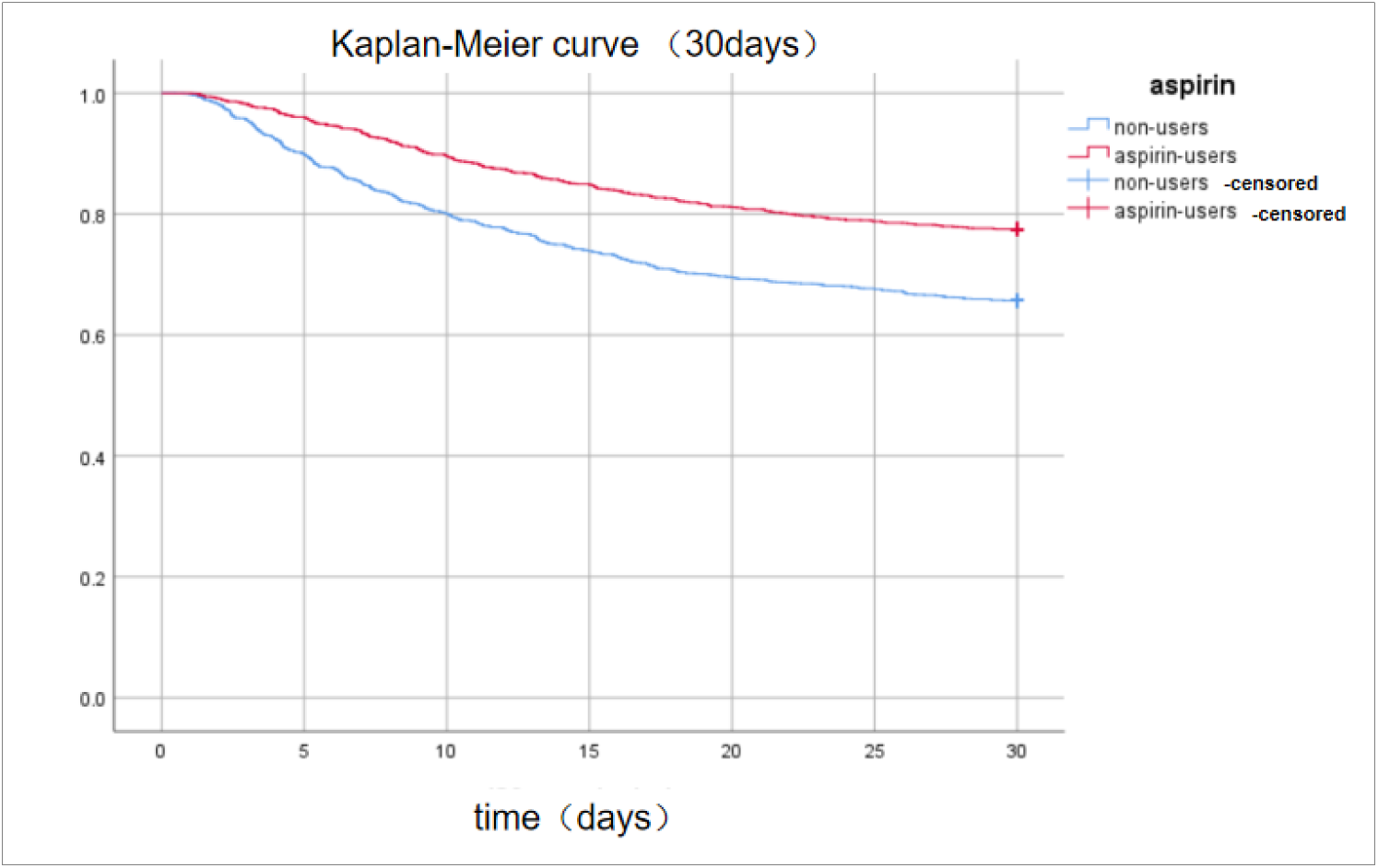
Kaplan-Meier survival curves between two groups indicated the 30-day mortality risk for the ARF patients. Non-aspirin users are represented by blue lines and aspirin users are represented by red lines

**FIGURE 3.**
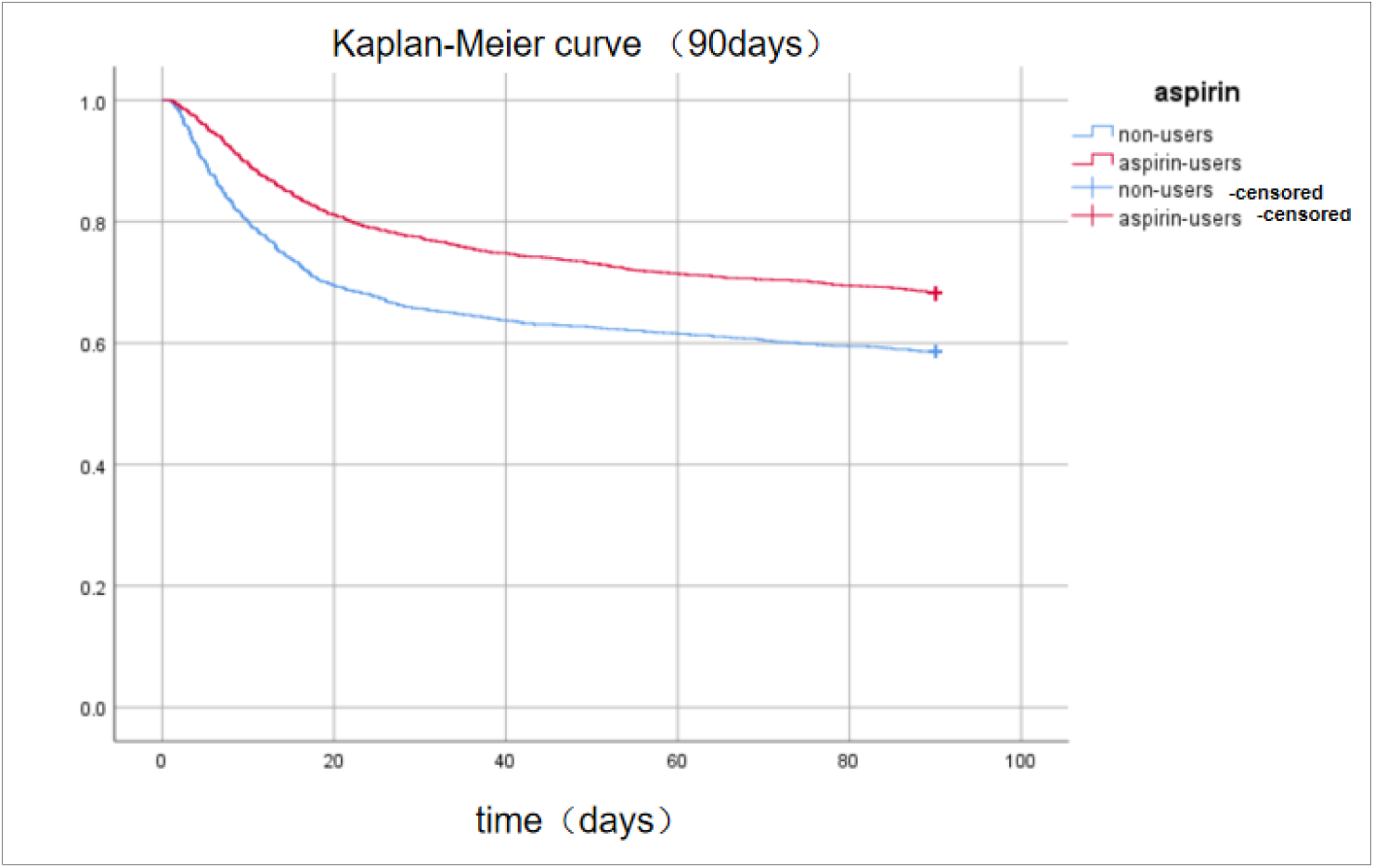
Kaplan-Meier survival curves between two groups indicated the 90-day mortality risk for the ARF patients. Non-aspirin users are represented by blue lines and aspirin users are represented by red lines.

### 4.3 Association of aspirin with composite outcomes

The group receiving aspirin had longer stays in the ICU and in the hospital, but notably shorter durations of mechanical ventilation (Table 3). This observation implies that although patients treated with aspirin might require more extended periods in critical care settings and hospitalization, their need for mechanical breathing support was reduced compared to those not receiving aspirin. This could potentially reflect improved respiratory function or faster recovery from respiratory failure in patients on aspirin therapy.

**TABLE 3.** Composite outcomes of the aspirin use group and the non-users group.

| Composite outcomes | Non-users | Users-Aspirin | p-value |
| --- | --- | --- | --- |
| Length of ICU stay,<br>median (inter-quartile<br>range) | 6.08[3.68,11.1775] | 6.93[3.87,11.95] | 0.004 |
| Length of hospital stay,<br>median (inter-quartile<br>range) | 11.78[6.96,19.20] | 14.13[8.77,22.78] | <0.001 |
| surviving patients | 1393 (69.5%) | 1594 (79.5%) | <0.001 |
| mechanical ventilation<br>length | 58.00[27.00,117.16] | 54.95[22.50,122.00<br>] | <0.001 |

### 4.4 Subgroup analyses

ARF patients were divided into different subgroups according to age, gender, race, cancer, co-morbidity, SOFA scores, vasoactive drug, ventilation, renal replacement therapy (RRT) and continue renal replacement therapy (CRRT).The impact of aspirin on 90-day mortality within the subgroups of ARF (Acute Renal Failure) patients was explored, and the findings were visually represented using a Forest Plot (depicted in Figure 4).The analysis revealed that aspirin was associated with a lower 90-day mortality rate in several subgroups, specifically: patients aged ≤60 years (HR: 0.621, 95% CI: 0.474 to 0.813),male patients (HR: 0.671, 95% CI: 0.585 to 0.769),non-Caucasian patients (HR: 0.553, 95% CI: 0.465 to 0.657),patients with sepsis (HR: 0.681, 95% CI: 0.611 to 0.759),patients with COPD(chronic obstructive pulmonary disease) (HR: 0.656, 95% CI: 0.551 to 0.781),hypertensive patients (HR: 0.664, 95% CI: 0.579 to 0.761),patients receiving vasopressor drugs (HR: 0.656, 95% CI: 0.581 to 0.741),patients on mechanical ventilation (HR: 0.686, 95% CI: 0.616 to 0.763),patients with a SOFA(sequential organ failure assessment) score>6 (HR: 0.662, 95% CI: 0.581 to 0.754).The Hazard Ratio (HR) between groups that did or did not use CRRT (Continuous Renal Replacement Therapy) showed a significant difference. We found that compared to patients who did not receive CRRT treatment (HR 0.660, p<0.001, 95% CI 0.589, 0.739), the benefits of using aspirin were limited in patients who received CRRT treatment (HR 0.838, p 0.174, 95% CI 0.65, 1.081). This could provide an explanation for the varied perspectives seen in earlier research concerning the effects of aspirin on ARF patients.

**FIGURE 4.**
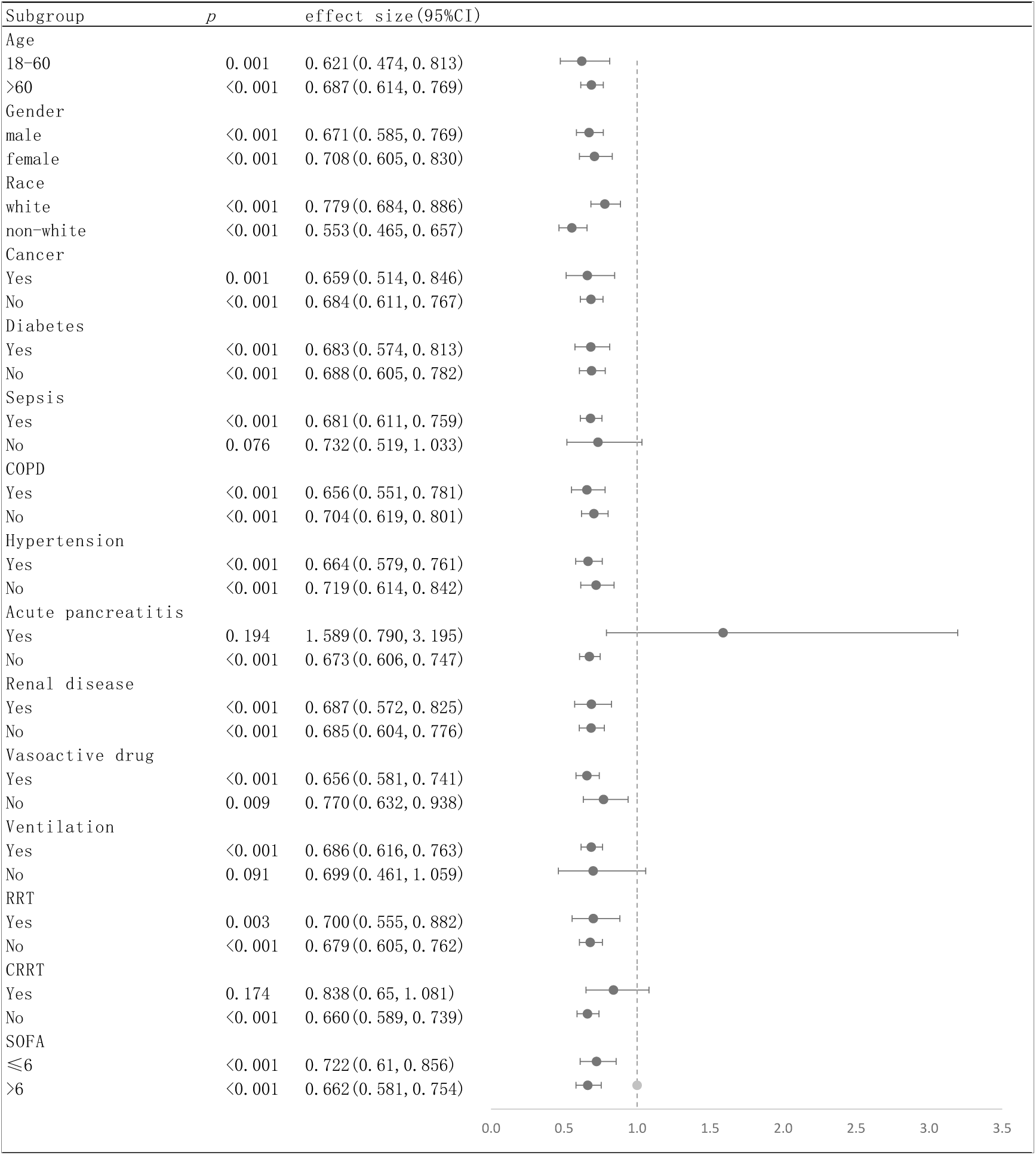
Subgroup analysis of the relationship between aspirin and 90-Day mortality, illustrated by a Forest Plot.

### 4.5 Dose of aspirin

To further investigate the optimal dose of aspirin for treatment, we analyzed the relationship between aspirin dosage and 90-day mortality using a Cox proportional hazards model. Patients diagnosed with ARF and treated with aspirin while ICU were categorized based on the dosage of aspirin they received. The study revealed that those in the high-dose group, who were prescribed more than 300 mg per day, faced a higher risk of mortality within 90 days as opposed to the low-dose group, which received 300 mg or less per day. The adjusted Hazard Ratio (HR) for the high-dose group was 1.332, with a 95% Confidence Interval (CI) of 1.094 to 1.623, and the statistical significance was confirmed with a p-value of 0.006, as presented in Table 4. The Cox proportional-hazards model, which provides a visual representation of these findings, is depicted in Figure 5.

**TABLE 4.** Survival outcomes of the high-dose aspirin group and the low-dose aspirin group

|  |  | No.<br>events/N<br>o. All<br>patients | Median survival<br>time (95%CI) | Log<br>rank<br>test<br>value | Hazard ratio<br>(95%CI) | Adjusted<br>hazard Ratio<br>(95%CI) |
| --- | --- | --- | --- | --- | --- | --- |
| 90-day<br>mortality | low-dose<br>aspirin | 497/1631 | 90.00(46.17,90.00) | 0.006 | HR=1.301<br>(1.078,1.569) | Adjusted<br>HR=1.332(1.094,1.623) |
|  | high-dose<br>aspirin | 140/373 | 10.48<br>(6.28,17.07) |  |  |  |

**FIGURE 5.**
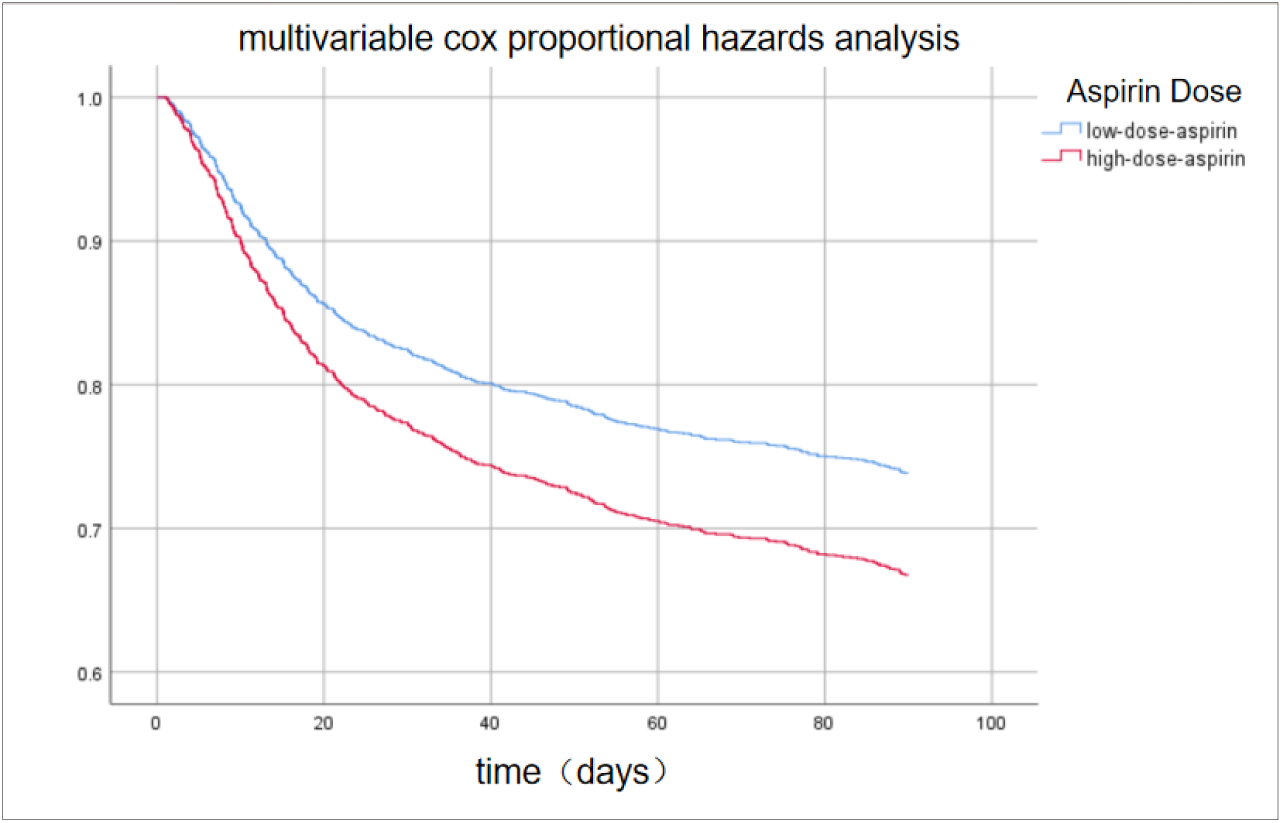
Cox proportional-hazards model of the dose of aspirin and 90-day mortality in ARF patients. Low-dose (300 mg/d) aspirin group is represented by blue lines and high-dose (>300 mg/d) aspirin group is represented by red lines.

## 5 Discussion

The therapeutic advantages of aspirin for patients with ARF are not well established, and this observational study aims to determine if aspirin has a positive impact on ARF patient outcomes. The findings suggest that aspirin therapy for patients with ARF in the ICU could potentially lower the risk of mortality at both the 90-day and 30-day marks,but may slightly prolong the duration of ICU and hospital stays.

Even with progress in the treatment of ARF, the death rate remains significantly high. This life-threatening condition is char acterised by alveolar damage, disruption of the endothelial-epithelial barrier, increased permeability, formation of pulmonary edema and decreased pulmonary gas exchange. It is reported that platelet activation plays a significant role in ARF, potentially mediating vascular leakage and increasing alveolar-capillary permeability due to compromised stability of endothelial cadherins^19^. Aspirin is considered a potential therapeutic option, not only for its antiplatelet effects but also for its anti-inflammatory properties. In the complex pathophysiology of ARF, which involves increased infammation and procoagulant factors and the destruction of the alveolar–capillary barrier, aspirin’s multifunctionality could ofer a novel therapeutic perspective^5,20,21^. And platelets, by expressing CD40, have the capacity to trigger the activation of additional platelets and endothelial cells. They also secrete a variety of signaling molecules such as cytokines and chemokines, which contribute to the inflammatory process. Furthermore, platelets are involved in the activation of the complement system, a component of the immune response, which enhances inflammation during ARF^19,22^.A study targeting sepsis-related respiratory failure patients has shown that pre-ICU exposure to aspirin is associated with a reduction in 28-day mortality, 60-day mortality, and in-hospital mortality in S-ARF (sepsis-associated acute respiratory failure) patients^23^. Aspirin was the first antiplatelet agent introduced into clinical practice and remains widely used today. By inhibiting COX-1, it blocks the synthesis of thromboxane A2, a potent platelet activator^24^.

The advantage of aspirin in ARF patients is probably due to its capacity to inhibit platelet function. Multiple relevant studies have indicated that antiplatelet medications can decrease the risk of death in ARF patients. Aspirin induced resolvin D1 has been shown to inhibit oxygen induced pulmonary edema, permeability, and inflammation in mice^25^. An observational study of 202 ICU-admitted ARDS patients demonstrated a significantly reduced ICU mortality rate in those using aspirin (odds ratio (OR) 0.38, 95% CI 0.15 to 0.96, P = 0.04)^26^. Aspirin is a potential anti-fibrotic drug for the treatment of patients infected with Severe Acute Respiratory Syndrome Coronavirus 2 (SARS-CoV-2)^27^.

The three fundamental mechanisms of ARF include microvascular dysfunction, inflammation, and metabolic reprogramming^19^. Aspirin possesses potent anti-inflammatory capabilities, which are due to its action in blocking the production of prostanoids and facilitating the generation of lipoxins and resolvins^28^. Previous studies have reported that aspirin triggered resolvin D1 (AT-RvD1) effectively downregulates the inflammatory response of lipopolysaccharide (LPS) - induced ARF mice, thereby reducing alveolar cells in endotoxin induced acute respiratory failure^25^. Resolvin D1(RvD1) is an anti-inflammatory bioactive compound that can downregulate NF-κB inflammatory signals , highlighting its potential as a therapeutic target in ARF^29,30^.

The efficacy of aspirin is contingent upon the dosage administered. At lower doses, aspirin acts irreversibly to inhibit cyclooxygenase-1 (COX-1), diminishing the capacity of platelets to synthesize and release the secondary aggregator thromboxane A2^16^. In contrast, higher doses of aspirin exert a broader inhibition, targeting both COX-1 and COX-2 enzymes, which enhances its anti-inflammatory activity^31^. In our research, it was observed that patients in the low-dose aspirin group had a higher 90-day survival rate compared to those in the high-dose aspirin group. This discrepancy might be attributed to the increased likelihood of side effects with higher doses of aspirin, including gastrointestinal bleeding, platelet dysfunction, allergic reactions, Reye’s syndrome, kidney damage, and drug interactions^32–37^. The subgroup analysis in our study showed that the benefit of using aspirin was even greater for patients with COPD (HR 0.656, 95% CI 0.551 to 0.781) and hypertension (HR 0.664, 95% CI 0.579 to 0.761) compared to non-COPD patients (HR 0.704, 95% CI 0.619 to 0.801) and non-hypertensive patients (HR 0.719, 95 CI 0.614 to 0.842) (Figure 4). A significant portion of ARF patients have a history of hypertension and COPD, both of which are harmful factors for cardiovascular-related mortality. Aspirin may help prevent ARF by exerting beneficial effects on the cardiovascular system.

Nonetheless, the limitations of the present study should be recognized. Firstly, we focused on the most critically ill patients, as the MIMIC database includes ICU admissions at the Beth Israel Deaconess Medical Center. Despite using propensity score matchiwng to minimize differences in baseline characteristics between the two groups, the complex nature of ICU environments means that the mortality of these patients may be influenced by other factors beyond aspirin use. Secondly, due to the observational nature of the study, the group assignments were not randomized. While propensity score matching was used to reduce bias between the aspirin and non-aspirin groups, there may still be residual confounding factors that affected the outcomes. Thirdly, this being a retrospective study, we cannot establish causality between aspirin use and ARF outcomes. Lastly, the long-term effects of aspirin on ARF outcomes could not be assessed due to the absence of long-term follow-up data. Therefore, further research, including animal experiments and additional prospective cohort studies, is warranted to investigate the relationship between aspirin use and the prognosis in ARF patients.

## 6 Conclusion

Aspirin may reduce the 30-day or 90-day mortality risk in ARF patients and also shorten the duration of mechanical ventilation. Aspirin could be an effective medication in the treatment of ARF patients. However, further studies are required to investigate the specific mechanism underlying aspirin benefiting ARF patients.

## Data Availability

All relevant data are within the manuscript and its Supporting Information files.

